# Agreement Between Actigraphy-Based Sleep Measures and Routine Nursing Sleep Assessments in Acute Psychiatric Inpatient Care

**DOI:** 10.64898/2026.09.08.26362317

**Authors:** Hong-Ming Chen, Yi-Lung Chen, Cheryl C-H Yang

## Abstract

**Introduction:** Sleep is routinely assessed by psychiatric mental health nurses in acute inpatient wards, where overnight observation forms part of everyday clinical care. These records are repeatedly collected, but observational assessment and activity-based monitoring capture different aspects of sleep and may not be interchangeable.

**Aim:** To examine the agreement between actigraphy-based total sleep time and routine nursing sleep assessments in an acute psychiatric inpatient ward.

**Method:** This cross-sectional observational study included 94 patients admitted to an acute psychiatric ward. Participants wore wrist actigraphy devices for up to seven available nights while sleep duration was concurrently documented by nursing staff as part of routine care. Pearson correlation coefficients assessed linear association and intraclass correlation coefficients (ICC) assessed agreement.

**Results:** Across all participants, nursing-assessed and actigraphy-based total sleep time showed a moderate linear association (Pearson r = 0.629) and good ICC (0.767). Bland– Altman analysis showed a mean bias of 0.20 hours, with 95% limits of agreement from -1.79 to 2.19 hours. Agreement varied across diagnostic groups and was strongest among patients with bipolar mania (r = 0.826; ICC = 0.886).

**Discussion:** Routine nursing sleep assessments showed little systematic bias relative to actigraphy at the group level, but wide limits of agreement indicate that the methods are not interchangeable for individual patients. Patient-reported sleep was not measured and should be included in future comparisons.

**Implications for Practice:** Routine nursing sleep records provide clinically meaningful longitudinal information. Actigraphy may serve as a selective adjunct when an independent estimate is needed, with implementation balanced against equipment, processing and workflow requirements.

**Accessible Summary:** *What is known on the subject?:*

- Sleep disturbance is common in acute psychiatric inpatient care, and psychiatric mental health nurses routinely observe and document sleep overnight.
- Nursing observation, patient report and actigraphy reflect different aspects of sleep; disagreement between methods can occur.

*What this paper adds?:*

- In 94 acute psychiatric inpatients, routine nursing sleep assessments showed moderate association and good agreement with actigraphy-based total sleep time.
- Agreement varied across diagnostic groups and was strongest in patients with bipolar mania.
- The findings support nursing sleep assessment as a clinically meaningful source of information while positioning actigraphy as an adjunct rather than a replacement.

*What are the implications for practice?:*

- Routine nursing sleep documentation may be useful for longitudinal monitoring in psychiatric inpatient care.
- Actigraphy may be useful when an independent or more detailed estimate of sleep is needed, but equipment, data-processing and workflow requirements should be considered.
- Patient-reported sleep was not collected; future work should compare all three perspectives rather than assuming that any single method represents sleep completely.

**Relevance to Mental Health Nursing:** Psychiatric mental health nurses generate repeated sleep observations as part of routine inpatient care. This study shows that those observations correspond meaningfully with an independent activity-based measure of sleep. The findings support continued clinical use of nursing sleep assessment and suggest a selective adjunctive role for actigraphy when additional objective information is required.

**Originality of the study:** Previous psychiatric inpatient work has compared nurse-reported sleep with wrist actigraphy. The present study extends this literature in a contemporary acute ward cohort of 94 patients by using repeated monitoring over up to seven available nights, examining three diagnostic groups, and characterizing both association/reliability and absolute agreement. Its contribution is therefore an updated real-world evaluation of routine nursing sleep assessment rather than the first comparison of nursing observation with actigraphy.

**Rigour of the study:** A pragmatic consecutive sample was observed under routine ward conditions. Sleep was measured concurrently by nursing assessment and wrist actigraphy over multiple nights. Pearson correlation quantified linear association, ICC summarized agreement/reliability, and Bland–Altman analysis quantified mean bias and limits of agreement. Reporting followed the STROBE statement.

**Significance of the study:** Routine nursing sleep documentation is inexpensive, repeatedly available and embedded in clinical workflow. Demonstrating meaningful agreement with an independent actigraphy-based measure supports its value as a source of longitudinal clinical information. Actigraphy may add information in selected situations, but should complement rather than displace nursing assessment.

## Introduction

Sleep assessment is part of everyday psychiatric mental health nursing in acute inpatient wards. Night staff repeatedly observe patients for safety and well-being, and these observations also generate longitudinal information about sleep. Nursing observation is a situated clinical practice: it occurs within ward routines and incorporates visible behaviour, environmental context and knowledge of the patient. Night-time observations can themselves disturb sleep, and the ward environment, observation frequency and staffing conditions may influence what is seen and recorded [8,13,14].

Nursing research has repeatedly identified sleep as an area in which assessment matters clinically but is poorly standardised. A review of sleep assessment in hospitalised patients concluded that standardised sleep measurement is not routinely undertaken in acute care and that no brief instrument has been established for everyday nursing use [17]. Subsequent nursing reviews report that nurses regard sleep as clinically important yet describe limited training, few structured assessment tools and inconsistent documentation [18], while evidence that nurse-delivered interventions can improve inpatient sleep continues to accumulate [19]. Routine nursing sleep records therefore occupy an unusual position in the discipline: they are generated at scale within existing workflows, yet their measurement properties have seldom been examined directly.

The clinical importance of sleep makes the quality of these routine observations consequential. Sleep and circadian disruption are common across schizophrenia, bipolar disorder and depressive disorders and are associated with psychiatric symptoms and illness course [1–4,12]. Yet sleep is not a single directly observed phenomenon. Patient report reflects perceived sleep, nursing assessment reflects intermittent clinical observation, and actigraphy estimates sleep-wake state from movement. Each approach therefore has distinct strengths and sources of error.

An earlier study in psychiatric inpatients compared nurse reports, patient sleep diaries and wrist actigraphy and found satisfactory agreement between nursing reports and actigraphy, whereas patient reports differed in other systematic ways [15]. This distinction matters here: discrepancies between patient-reported and nursing-recorded sleep are clinically important, but patient-reported sleep was not collected in this study. The present question is narrower—how closely routine nursing estimates of sleep duration correspond with an independent actigraphy-based estimate under real-world acute ward conditions.

Actigraphy offers low-burden longitudinal monitoring and is less resource intensive than polysomnography, but it is not cost-free and does not directly measure sleep. Activity-based algorithms have high sensitivity for sleep but lower specificity for quiet wakefulness [5–7,9]. Compared with routine nursing documentation, implementation also requires devices, data extraction and processing, staff familiarity, maintenance procedures and attention to adherence. Its practical value therefore depends not only on measurement performance but also on whether it adds useful information beyond observations already embedded in nursing care [16].

Evidence directly evaluating routine nursing sleep assessment against actigraphy in acute psychiatric wards remains limited. Clarifying this relationship informs how nurses should interpret routinely recorded sleep duration and helps define when an additional wearable measure may be justified. We therefore examined agreement between actigraphy-based total sleep time and routine nursing sleep assessments in 94 acute psychiatric inpatients, including exploratory diagnostic subgroup analyses.

## Methods

### Participants

This cross-sectional observational study was conducted at a psychiatric acute ward in southern Taiwan between September 2019 and March 2023. This study was reported in accordance with the Strengthening the Reporting of Observational Studies in Epidemiology (STROBE) statement. A total of 94 patients aged 15–80 years were enrolled in the study from the psychiatric acute ward. Participants were screened upon admission and became eligible for enrollment once their psychiatric condition had sufficiently improved to permit informed consent.

After written informed consent was obtained, a trained research assistant explained the study procedures and assisted participants with placement of the actigraphy device. Participants were instructed to wear the wrist-worn actigraphy device during the intended monitoring period and contributed up to seven available nightly observations. Nursing sleep assessments were recorded concurrently as part of routine ward care. Nursing and actigraphy measurements were aligned by observation night, and agreement analyses were based on paired measurements from the same nights; observations were not necessarily obtained on consecutive calendar days. During routine evening medication administration (approximately 20:30), nursing staff informally verified continued device use as part of routine ward care. No formal records regarding refusal rates, device loss, adherence rates, or device damage were maintained. This should be considered a limitation of the present study. Participants were categorized into three diagnostic groups: schizophrenia, bipolar mania, and depressive or anxious disorders (the depressive/anxiety disorders group).

### Patient Inclusion/exclusion

A pragmatic consecutive sampling approach was used, whereby all eligible patients admitted during the study period were invited to participate. Given the exploratory and real-world nature of this study, a formal a priori sample size calculation was not performed. Instead, the sample size was determined pragmatically based on consecutive admissions during the study period, which is consistent with previous actigraphy studies conducted in inpatient clinical settings. We excluded individuals under compulsory admission, those with obvious withdrawal symptoms from alcohol or other substances, patients showing clear signs of delirium, and those with intellectual disability or dementia.

### Validity and reliability of the actigraphy

Actigraphy provides an activity-based estimate of sleep–wake patterns and has been widely used in clinical sleep research. Compared with polysomnography, actigraphy demonstrates high sensitivity but lower specificity, particularly in distinguishing quiet wakefulness from sleep [9]. Sleep-related data were collected using a wrist-worn accelerometer (XA-5 model), which has been used previously in clinical sleep research [11]. This accelerometer was designed to be worn around the wrist, enabling continuous actigraphic recording during the intended monitoring period.

### Statistical analysis

**Statistical analyses were performed to evaluate the relationship and agreement between actigraphy-based sleep estimates and routine nursing sleep assessments. Agreement analyses used paired nursing and actigraphy measurements from the same observation nights. Among nights with observations available from both sources and verifiable timestamps, nursing and actigraphy records were aligned to the same calendar night. Pearson correlation coefficients were used to assess linear associations, and intraclass correlation coefficients (ICC) were calculated to assess agreement [10]. ICC estimates were based on a two-way random-effects model using an absolute agreement definition and are reported as average-measures estimates (ICC(2,k)). Bland– Altman analysis was additionally used to quantify absolute agreement for total sleep time (TST), with the difference defined as actigraphy minus nursing assessment. Mean bias and 95% limits of agreement (mean difference ± 1.96 SD) were calculated. Subgroup analyses were conducted across diagnostic categories. A p value of < 0.05 was considered statistically significant.**

## Results

### Participant characteristics

**Of the 96 patients initially screened, 94 completed the study procedures and were included in the final analysis. Two participants were excluded because they were enrolled outside the approved study recruitment period and therefore did not meet protocol requirements. The sample consisted of 58 men (61.7%) and 36 women (38.3%), with a mean age of 43.10 years (SD = 15.0). Participants included individuals with schizophrenia (n = 32, 34.0%), bipolar mania (n = 31, 33.0%), and depressive or anxiety disorders (n = 31, 33.0%). The median admission duration was 23.0 days (IQR 16.0–32.8), and participants were enrolled a median of 4.0 days (IQR 2.0–7.0) after admission. Participants contributed 5–7 available nightly observations; 86/94 (91.5%) had seven available observations. The observation series spanned a median of 8 days (IQR 7–10; range 5–19), and 39/94 (41.5%) series were obtained on consecutive calendar days.**

### Diagnostic subgroup

Clinical characteristics across diagnostic groups are presented in Table 2. No significant differences were observed among diagnostic groups in age, sex distribution, or admission duration. However, the median enrollment day differed significantly across groups (p = 0.013), with patients in the schizophrenia group enrolling later after admission than those in the bipolar mania and depressive/anxiety disorders groups.

**Table 1.** Participant Characteristics (N = 94)

| Characteristic | Value |
| --- | --- |
| Age, mean (SD), years | 43.10 (15.0) |
| ***Sex, n (%)*** |  |
| Male | 58 (61.7) |
| Female | 36 (38.3) |
| ***Primary diagnosis, n (%)*** |  |
| Schizophrenia spectrum disorders | 32 (34.0) |
| Bipolar mania | 31 (33.0) |
| Depressive/anxiety disorders | 31 (33.0) |
| Admission length, median (IQR), days | 23.0 (16.0–32.8) |
| Enrollment day after admission, median (IQR), days | 4.0 (2.0–7.0) |
| Available nightly observations, median (IQR) [range] | 7 (7–7) [5–7] |
| Participants with 7 available nightly observations, n (%) | 86 (91.5) |
| Calendar span of observations, median (IQR) [range], days | 8 (7–10) [5–19] |
| Consecutive calendar-day observation series, n (%) | 39 (41.5) |

**Table 2.**
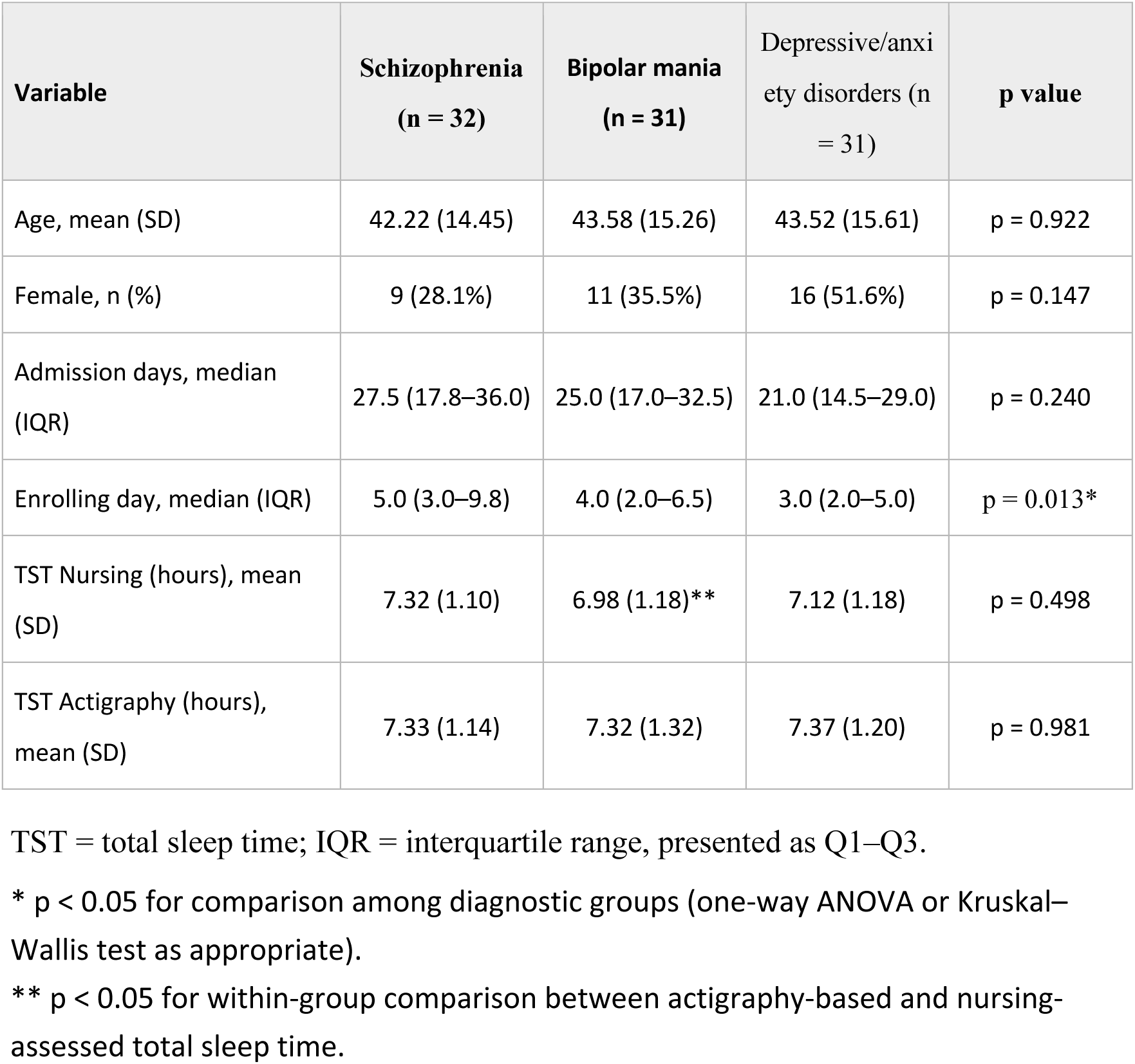
Clinical and Sleep Assessment Characteristics by Diagnostic Group.

**Total sleep time (TST)** estimated by nursing observation did not differ significantly across diagnostic groups overall. Similarly, actigraphy-derived TST showed no significant differences between groups. However, within the bipolar mania subgroup, actigraphy-based sleep estimates were significantly longer than nursing-assessed sleep duration (p < 0.05).

### Agreement between actigraphy and nursing sleep assessments

Agreement between actigraphy-based sleep measures and routine nursing sleep assessments is presented in Table 3. Across all participants, the Pearson correlation coefficient between the two methods was 0.629, indicating a moderate association. The intraclass correlation coefficient (ICC) was 0.767 (95% CI, 0.65 to 0.84), suggesting good agreement between the two measurement approaches.

**Table 3.**
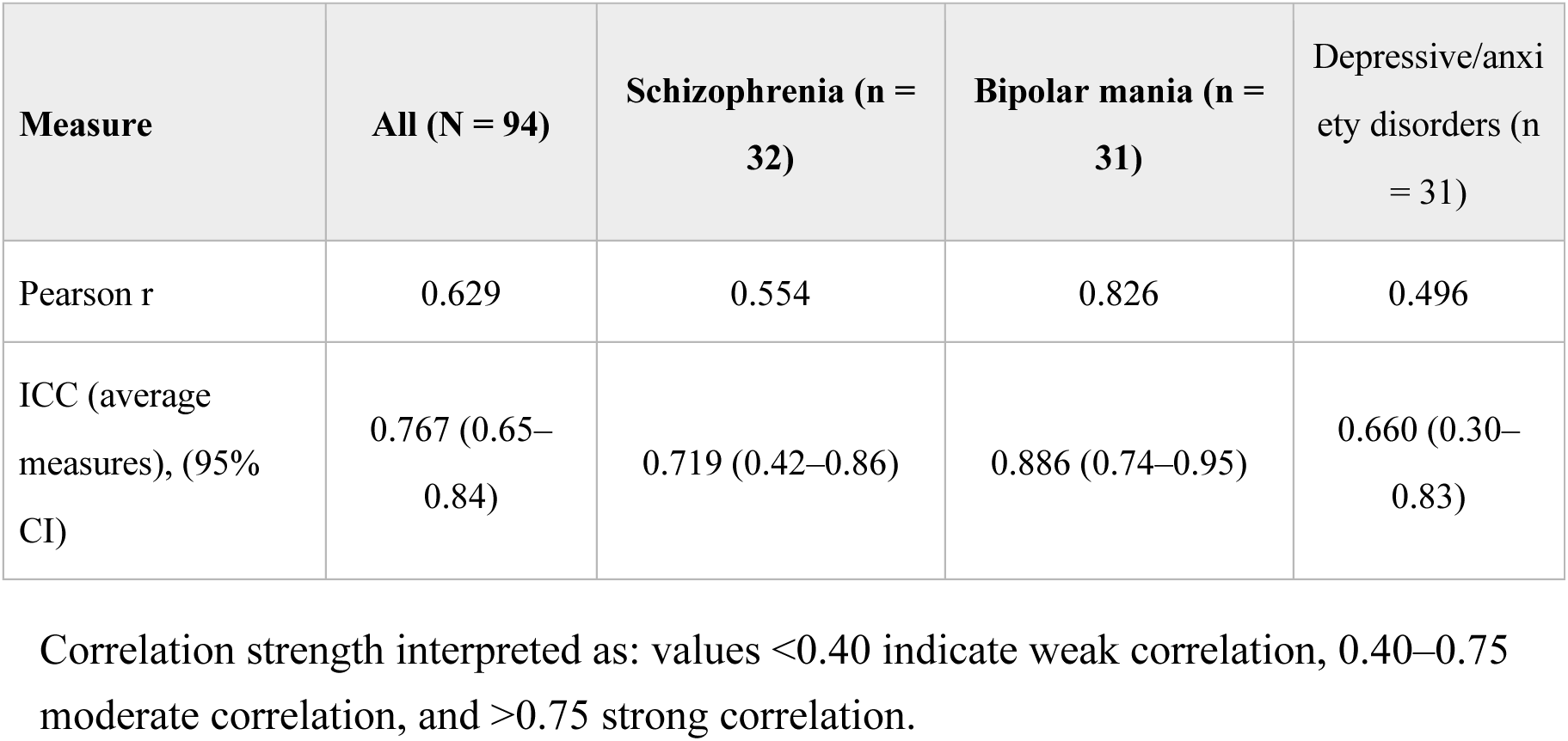
Agreement Between Actigraphy and Nursing Sleep Assessments.

Subgroup analyses demonstrated variation in the strength of agreement between methods. The bipolar mania subgroup showed the strongest association (Pearson r = 0.826; ICC = 0.886), whereas the depressive/anxiety disorders subgroup demonstrated a comparatively lower correlation (Pearson r = 0.496; ICC = 0.660). All correlations were statistically significant.

Bland–Altman analysis showed little systematic difference between methods. The mean bias (actigraphy minus nursing assessment) was 0.20 hours (95% CI, -0.01 to 0.41), with 95% limits of agreement from -1.79 to 2.19 hours (Figure 1). Thus, group-level estimates were similar on average, while individual-level differences could approach approximately two hours in either direction.

**Figure 1.**
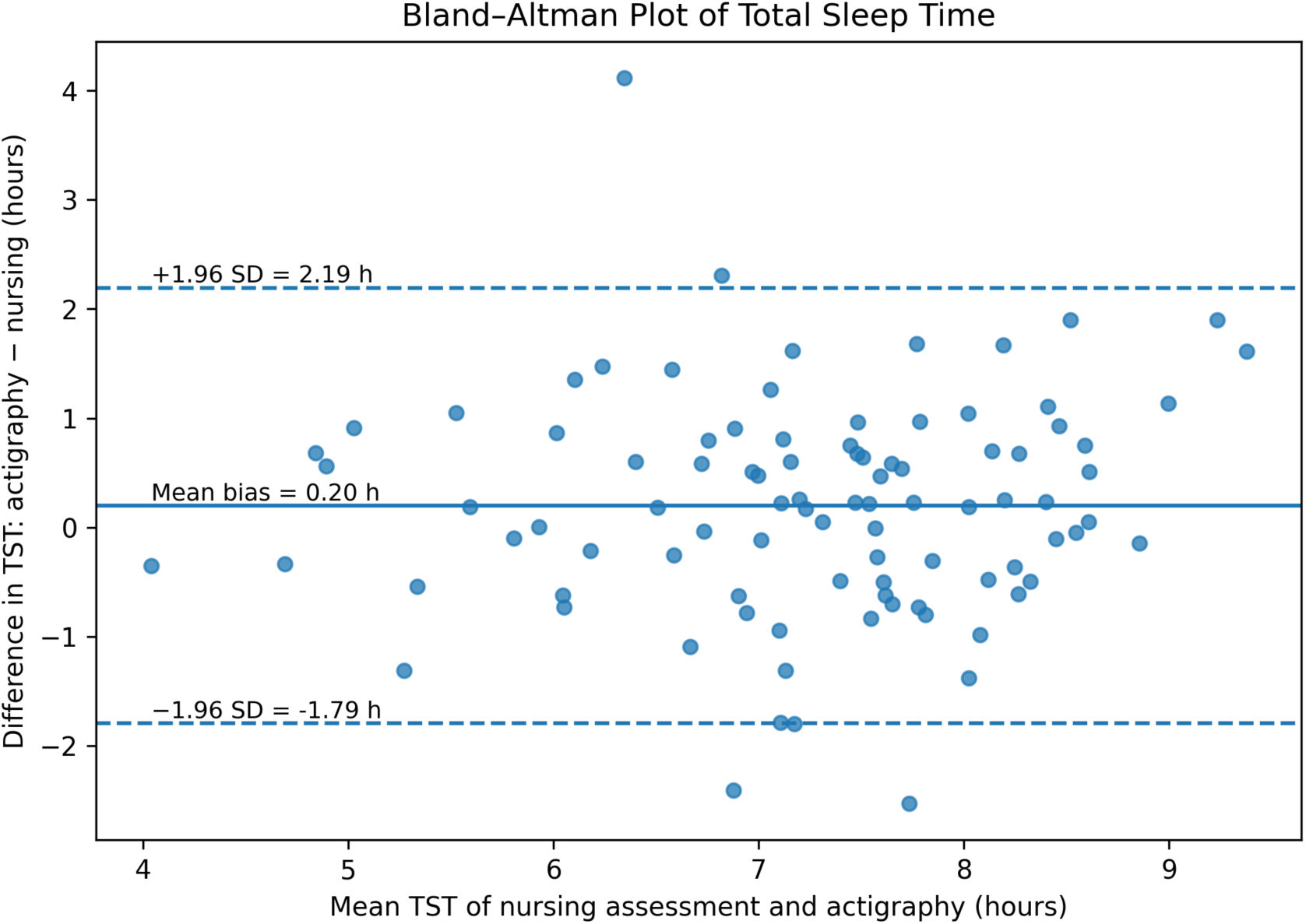
Bland–Altman plot comparing actigraphy-based and nursing-assessed total sleep time. The solid line indicates mean bias; dashed lines indicate the 95% limits of agreement.

## Discussion

### Routine nursing sleep assessment as a measurement source

The principal finding is that routine nursing assessment of total sleep time corresponded meaningfully with an independent actigraphy-based estimate in an acute psychiatric inpatient ward. Across all participants, the association was moderate (r = 0.629) and agreement was good (ICC = 0.767). These findings support the clinical information value of sleep observations already generated by psychiatric mental health nurses during routine care. They do not establish that nursing assessment and actigraphy are interchangeable.

The two methods observe sleep differently. Nursing assessment is based on intermittent observation interpreted within the clinical context of the ward, whereas actigraphy infers sleep and wake from motor activity. Nurses may miss periods of quiet wakefulness between observations; conversely, actigraphy can classify motionless wakefulness as sleep and cannot capture the behavioural context available to nursing staff [9]. Agreement therefore indicates convergence between two different measurement perspectives rather than proof that either method represents a definitive ground truth.

The findings are consistent with earlier psychiatric inpatient work showing satisfactory agreement between nursing reports and actigraphy and demonstrating that patient self-report can differ systematically from both [15]. Patient-reported sleep was not collected here. Accordingly, this study cannot resolve discrepancies between patients’ sleep experience and nursing documentation. Future studies should measure patient report, nursing assessment and actigraphy concurrently because each may contribute non-redundant information.

### Diagnostic subgroup findings

Agreement varied across diagnostic groups and was strongest among patients with bipolar mania (r = 0.826; ICC = 0.886). At the whole-sample level, Bland–Altman analysis further clarified the nature of agreement: mean bias was small (+0.20 hours), but the 95% limits of agreement were relatively wide (-1.79 to +2.19 hours). Therefore, nursing assessment and actigraphy produced similar average TST at the group level but should not be treated as interchangeable measurements for an individual patient.

The depressive/anxiety disorders subgroup showed the lowest agreement, while the schizophrenia subgroup showed moderate agreement. These subgroup findings are exploratory because sample sizes were small and the study was not powered for diagnostic comparisons. The data do not establish diagnosis-specific measurement rules.

### Implications for psychiatric mental health nursing

The most immediate implication is not that every psychiatric inpatient requires a wearable device. Routine nursing sleep documentation is already embedded in care, requires no additional sensor purchase and provides repeated observations across hospitalization. The present findings suggest that these records contain clinically meaningful information and should not be treated simply as an inferior substitute for device-based measurement.

A second implication concerns how nursing sleep data are valued as a data source. Nursing documentation already generates a large, longitudinal and routinely collected record of inpatient sleep, and secondary analysis of such nursing data is increasingly recognised as a means of advancing nursing science, quality improvement and service planning [20]. The present findings indicate that this existing record is not merely a low-quality proxy awaiting replacement by sensors: at the group level it tracked an independently generated activity-based estimate closely. Where analyses rest on aggregated or repeated observations, such as ward-level monitoring, quality improvement work, or research on sleep across an admission, routine nursing records provide coverage that a wearable deployment could not match without substantial new investment, because routine night-time observation is already embedded in inpatient care. The wide limits of agreement mean that this argument applies to patterns across nights rather than to the absolute sleep duration recorded for one patient on one night.

Framed this way, the practical question for a ward is not whether to replace nursing observation with devices, but whether the existing nursing record is documented consistently enough to support that role. Improving the consistency and structure of routine sleep documentation may generate more usable information, at lower cost, than adding sensors to a ward whose records already perform adequately. Conversely, where staffing levels or documentation quality cannot sustain reliable night-time observation, device-based measurement becomes correspondingly more valuable, and the case for investing in it strengthens.

Within this framing, actigraphy is best considered a selective adjunct. It can provide continuous, independently generated activity-based information across multiple nights and may be useful when greater temporal resolution or an independent estimate is needed. These potential benefits must be weighed against device acquisition and replacement costs, data-processing infrastructure, staff training and workflow, maintenance, adherence monitoring and the possibility of device loss or damage. Actigraphy is less resource intensive than polysomnography but more resource intensive than routine documentation or paper sleep logs [16].

This adjunctive framing preserves the clinical role of nursing observation. Psychiatric nursing observation produces contextual knowledge about behaviour and safety within the ward [13]. Digital measures can add another perspective, but they do not reproduce that situated clinical judgement. The practical question is when an additional measurement layer changes interpretation or care sufficiently to justify its burden.

### Strengths and limitations

The study was conducted under routine acute ward conditions, used concurrent nursing and actigraphy measurements over several days and included patients from three common diagnostic groups. This naturalistic design increases relevance to everyday psychiatric mental health nursing.

Several limitations warrant consideration. Enrollment occurred only after patients had improved sufficiently to provide informed consent, creating selection bias and limiting generalizability to the most acutely unwell phase. Monitoring was concentrated relatively early in hospitalization. Nightly observations were not always obtained on consecutive calendar days, and observation spans therefore varied among participants. However, nursing and actigraphy measurements were aligned by observation night for paired comparisons. Formal adherence, refusal, device-loss and device-damage data were not systematically recorded, and standardized symptom severity measures were unavailable.

Most importantly, the study did not collect patient-reported sleep or polysomnography. Actigraphy is therefore an independent comparator rather than a gold standard, and the results should not be interpreted as establishing the accuracy of either nursing assessment or actigraphy against physiologically measured sleep. The lack of patient-reported sleep also prevents direct evaluation of the clinically familiar situation in which patients describe poor sleep despite apparently adequate nursing-recorded duration.

### Future research

Future studies should compare patient report, nursing assessment and actigraphy concurrently and examine how disagreement among these perspectives relates to symptoms, treatment changes and patient experience. Larger samples could test whether diagnostic or behavioural characteristics systematically alter agreement. Implementation studies should also report adherence, device loss, staff workload and costs so that the incremental value of actigraphy can be judged against routine nursing assessment.

A second opportunity is to evaluate routine nursing sleep records longitudinally. Because these observations are repeatedly collected as part of ordinary care, nursing-recorded sleep measures may support larger pragmatic studies of inpatient sleep patterns without requiring continuous wearable deployment in every patient. Such use will require attention to documentation practices, missingness and variation across wards and institutions.

## Conclusion

Routine nursing assessments of total sleep time showed meaningful agreement with actigraphy-based estimates in acute psychiatric inpatient care. The findings support nursing sleep documentation as a clinically informative measurement source while showing that nursing observation and actigraphy should not be considered interchangeable. Actigraphy is best positioned as a selective adjunct when an independent or more detailed activity-based estimate is needed. Future work should incorporate patient-reported sleep and evaluate whether the additional information provided by wearable monitoring justifies its implementation burden.

## Data Availability Statement

The data that support the findings of this study are available on request from the corresponding author. The data are not publicly available due to privacy or ethical restrictions.

